# Genomic Feature Transformation for Multi-Omics Integration in Clinical Prediction Models

**DOI:** 10.64898/2026.03.09.26347570

**Authors:** Barry Ryan, Christian W. Thorball, Mariam Ait Oumelloul, Roger Kouyos, Philip E. Tarr, Jacques Fellay

## Abstract

Multi-omics integration in clinical prediction models relies on capturing cross-modal interactions to improve prediction beyond single-omics baselines. Incorporating genomics into such models remains non-trivial: genomic data is extremely high-dimensional, sparse in per-variant information content, and typically requires far larger cohorts than other omics layers to yield informative features. As a result, methods for integrating genomics with complementary omics data remain poorly standardised. Using the All of Us (AoU) and Swiss HIV Cohort Study (SHCS) datasets, we trained linear and deep-learning classifiers in single-omic and multi-omic settings to predict Coronary Artery Disease (CAD) and Chronic Kidney Disease (CKD), two conditions with established genetic and multi-omic associations. Genomics data was represented in four ways: (i) raw SNP genotype matrices, (ii) Principal Component Analysis (PCA), (iii) Polygenic Risk Scores (PRS), and (iv) AlphaGenome-derived gene-level functional impact scores. Each representation was evaluated individually and integrated with complementary omics via feature concatenation or a deep-learning encoder. Performance was estimated using nested three-fold cross-validation, with identical patient splits held constant across all genomic representations and their combinations; we report mean macro-averaged F1 score and standard deviation, with paired significance testing against raw SNP matrices. Genomic data alone was consistently outperformed by other individual omics layers. When integrated into multi-omics models, raw SNP matrices and PCA representations frequently degraded performance relative to genomics-free multi-omics models, whereas PRS matched or improved performance across most cohort-disease combinations and significantly outperformed raw SNPs for CAD prediction in the SHCS. Simple feature concatenation performed as well as, or better than, more complex encoder architectures, suggesting that genomic signal is captured in a largely additive rather than complementary manner at current sample sizes. These findings indicate that biologically informed transformations such as PRS, applied prior to integration, offer a practical strategy for incorporating genomics into multi-omics prediction models without requiring larger cohorts.

## Introduction

Personalised medicine promises to transform healthcare by enabling accurate prediction of disease risk, subtype classification, and optimal treatment selection for individual patients. Central to this paradigm is the development of AI-driven multi-omics models, which integrate diverse molecular data layers, such as genomics, proteomics, and transcriptomics, to build comprehensive patient representations [Ryan et al., 2024, Li et al., 2022, Wang et al., 2021].

Genomic features present unique challenges for integration in deep learning multi-omics frameworks. They are extremely high-dimensional, routinely comprising millions of Single Nucleotide Polymorphisms (SNPs), while also exhibiting low information density. Genomic data requires much larger cohorts than other omics layers, such as transcriptomics or proteomics. Thus, in integrative multi-omics models this disparity in availability limits the statistical power that genomics can contribute. Together, these limitations complicate both model training and cross-modal integration. As a result, methods for incorporating genomics alongside other omics layers remain poorly standardised, representing a significant gap in current AI-based multi-omics prediction models.

To keep analyses tractable and reduce redundancy, current multi-omics methods perform aggressive dimensionality reduction (e.g., Principal Component Analysis (PCA)) or restrict the number of SNPs to a subset without any biological rationale (e.g., 1000 SNPs of highest variance in the dataset) [Ryan et al., 2024, Ma et al., 2025, Wang et al., 2021, Li et al., 2022].

In this study, we evaluate several genomic feature transformation strategies within a multi-omics framework, assessing their impact on the prediction of two clinically important conditions: Coronary Artery Disease (CAD) and Chronic Kidney Disease (CKD). Both conditions are highly prevalent, have well-established genetic components, and have been associated with multiple omics layers, making them well-suited benchmarks for evaluating genomic data integration approaches [Khan et al., 2022, Patel et al., 2023].

Specifically, we investigate two complementary transformation approaches: the use of Genome Wide Association Studies (GWAS) summary statistics to construct compact, trait-specific Polygenic Risk Scores (PRS) features, and the application of foundation DNA models, which have demonstrated promise in capturing population-level patterns in genomic sequences [Avsec et al., 2026, 2021]. Notably, recent models such as AlphaGenome highlight the potential of such approaches to predict multiple omics layers directly from genomic data alone, offering a powerful avenue for biologically informed feature extraction.

We assess the impact of each transformation strategy on multi-omics classification performance using the All of Us (AoU) database, and independently validate our findings in the Swiss HIV Cohort Study (SHCS). Taken together, this work aims to establish best practices for genomic feature transformation, with the broader goal of improving genomic integration within AI-based multi-omics frameworks and advancing the accuracy and clinical utility of personalised medicine predictions.

## Methods

### Study Design

Table 1 summarises sample availability, cohort sizes, and omics feature counts. In the AoU dataset, all participants had genomics, transcriptomics, and proteomics data available. In the SHCS, all individuals had genomic data; the CAD subset also had proteomic profiles, and the CKD subset had metabolomics profiles. Where possible, three to one matching was performed in the AoU dataset due to a larger sample availability, however, only approximately one to one patient matching was achievable in the SHCS. In each case, omics samples were taken post diagnosis, making predictions in this setting diagnostic.

**Table 1.** Dataset overview and feature counts per condition in AoU and SHCS.

| Database | Cohort | Participant Counts |  | OtherOmics |  |  | Genomic Data |  |  |  |
| --- | --- | --- | --- | --- | --- | --- | --- | --- | --- | --- |
|  |  |  |  | Transcriptomics | Metabolomics | Proteomics | Pruned SNPs | PCA | PRS | Alpha Genome |
| AoU | CKD | Case | 328 | 32778 | NA | 5415 | 50000 | 1131 | 1799 | 58829 |
|  |  | Control | 973 |  |  |  |  |  |  |  |
|  | CAD | Case | 612 | 32778 | NA | 5415 | 50000 | 1958 | 1799 | 59307 |
|  |  | Control | 1676 |  |  |  |  |  |  |  |
| SHCS | CKD | Case | 166 | NA | 1930 | NA | 25605 | 285 | 3096 | 24310 |
|  |  | Control | 166 |  |  |  |  |  |  |  |
|  | CAD | Case | 431 | NA | NA | 766 | 25315 | 725 | 3096 | 47829 |
|  |  | Control | 418 |  |  |  |  |  |  |  |

In the AoU dataset, CAD cases were defined as participants with at least one ICD-10 code in the range I20–I25, while CKD cases were defined as those with an ICD-10 code of N18 or N19.

In the SHCS, CAD cases were defined as a clinical event of myocardial infarction, coronary angioplasty/stenting, or coronary artery bypass grafting. This definition is in accordance with standard definitions, such as those used by the Data Collection on Adverse events of Anti-HIV Drugs (D:A:D) study and the MONICA Project of the World Health Organization, as reported previously [Schoepf et al., 2021].

CKD cases were defined as a confirmed (over *≥* 3 months apart) decrease in eGFR from a baseline value *≥* 90 to *≤* 60*mL/minute/*1.73*m*^2^, in line with the Kidney Disease–Improving Global Outcomes (KDIGO) algorithm and previous large-scale investigations on CKD in PWH [Mocroft et al., 2016]. Only eGFR measurements per protocol were used, to avoid confounding by short term changes in kidney function due to, for example, transient use of kidney-toxic medication or dehydration. As a measure of kidney function, we calculated eGFR using the established Chronic Kidney Disease Epidemiology Collaboration equation, which has been validated in PWH [Bonjoch et al., 2010, Gagneux-Brunon et al., 2013, Levey et al., 2009, Cristelli et al., 2017].

### Genomic Data Transformations

Genomic data transformations investigated in this study are summarised in Figure 1.These transformations were performed seperately in both AoU and SHCS databases. In the AoU, whole genome sequencing has been performed on 553949 participants resulting in 132 million SNPs. In the SHCS genotyping arrays had been performed on 8228 PWH in the SHCS, revealing ∼86 million individual SNPs. Using PLINK version 1.9 by Chang et al. [2015], ∼6,5 million from the AoU and ∼3,8 million from the SHCS were extracted which had a minimum allele frequency of 5%, a Hardy-Weinberg equilibrium greater than 1*e*^−6^ and missingness below 1%. These SNPs were used for downstream analysis to generate PRS and to calculate gene-level impact scores using AlphaGenome.

**Figure 1.**
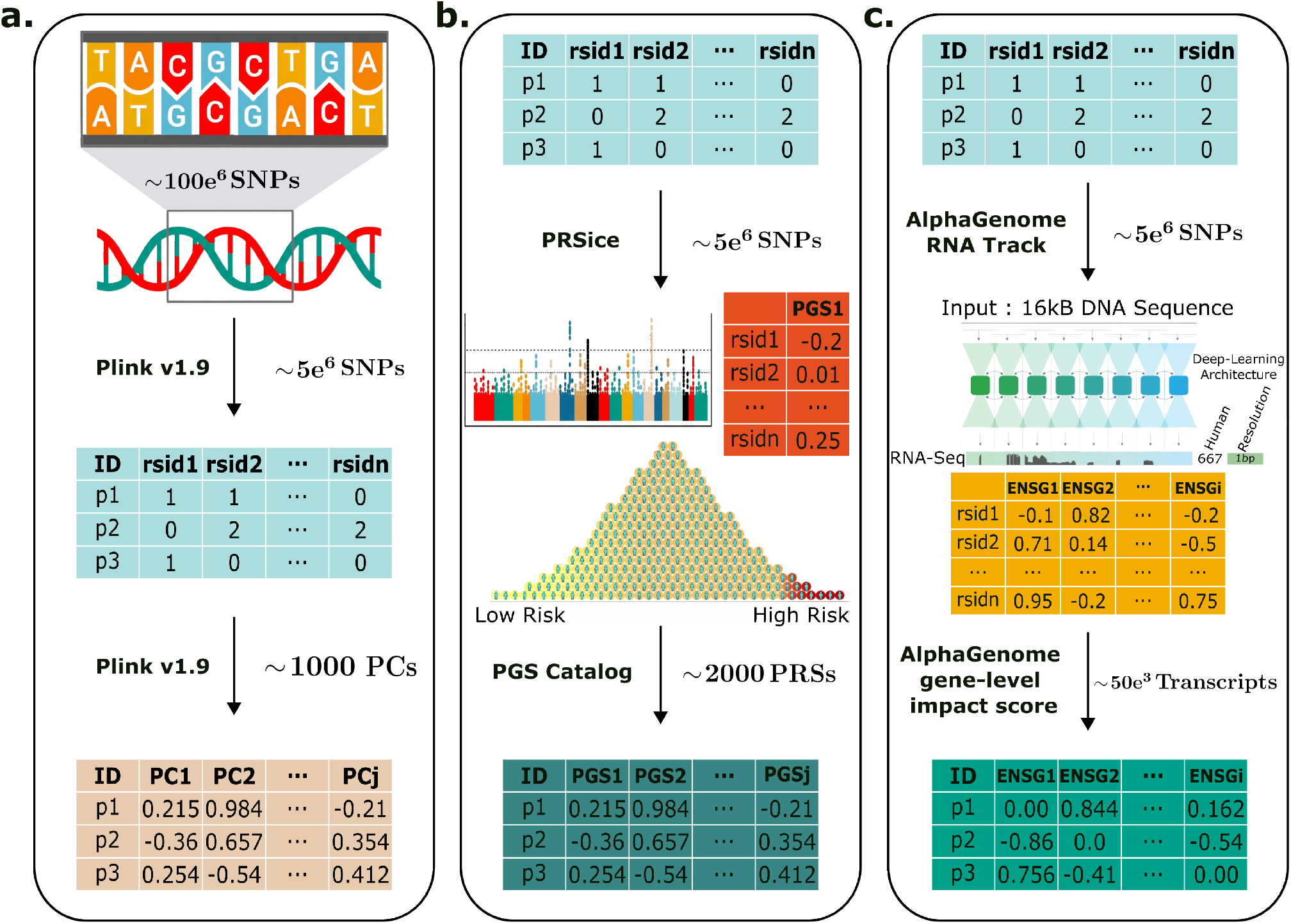
Summary of genomics data transformations. (a) PLINK v1.9 was used to extract ∼ 5 million SNPs, which were then pruned to the most variable within the CAD and CKD cohorts. The pruned SNPs were used to compute PCA and were also supplied directly as inputs to the classification models. (b) On the un-pruned SNPs, PRSice was used to compute PRS for all phenotypes and GWAS weights available in the PGS Catalog. (c) The un-pruned SNPs were also processed with AlphaGenome to calculate gene-level impact scores for each variant within a 16 kb window in two tissues, each relevant to one comorbidity

SNPs in linkage disequilibrium were pruned to remove near-redundant variants, preserving local haplotype structure and marker diversity while reducing redundancy for downstream modelling. Following pruning, 25,315 SNPs were retained in the CAD cohort and 25,605 in the CKD cohort within the SHCS. In the AoU datasets, over 250k SNPs remained across both cohorts. To ensure computational tractability, these were further reduced to the 50k most variable SNPs in each cohort. The resulting pruned variant sets served as both raw inputs to the classification models and as the basis for PCA-based dimensionality reduction.

As shown in Figure 1b PRSs were calculated using PRSice (version 2.3.3) software on all un-pruned ∼5 million SNPs. PRSs were calculated on all available phenotypes in the PGS Catalog, with information on included variants in each score and their weights also downloaded from the PGS Catalog [Lambert et al., 2021]. In the SHCSs, every available phenotype per individual was used in downstream analyses, as none of the PRS from PGS Catalog have been trained, tuned, or validated on the SHCS. However, PRS scores which used the AoU dataset to train the score were removed, likely resulting in the smaller set of PRS scores in the AoU cohorts as per Table 1.

Figure 1c demonstrates the input of SNPs into AlphaGenome [Avsec et al., 2026], which includes a built-in function to estimate the effect of a single variant within a genomic region. Using the RNA-seq track, we calculated the predicted transcriptional impact of each SNP on its corresponding gene within a 16 kb aggregation window, separately for two tissues: UBERON:0001621 (coronary artery) for CAD and UBERON:0002113 (kidney) for CKD. For each individual, we then computed a gene-level functional impact score by applying a weighted sum across all SNPs mapping to that gene, using the AlphaGenome-derived scores as weights in place of conventional GWAS effect sizes. This was implemented using PRSice’s weighted summation framework, weighted by allelic dosage. The resulting scores are structurally analogous to a PRS but reflect predicted functional consequence rather than statistical disease association.

### Models and Multi-Omics Integration

Genomics data transformations were evaluated for their ability to classify CAD and CKD, both separately and within a combined multi-omics setting. Each genomic data type and the complementary omics were passed into a linear model and a deep-learning model for classification. The linear classification model was a lasso logistic regression model implemented using the scikit-learn Python package. The deep-learning model consisted of a two layer Multi-layer Perceptron (MLP) implemented using PyTorch in Python. ReLU activation and dropout were applied with Adam optimisation and a cross entropy loss function.

Each genomic data transformation was also paired with available complementary omics data using two integration strategies. The first, feature-level concatenation, appended each genomic feature set with omics data. The resulting combined feature matrices were then passed to the same logistic regression and MLP models described previously.

Each genomic data transformation was also integrated with omics data using a deep-learning multimodal encoder, a common approach in multi-omics modelling [Ryan et al., 2024, Li et al., 2022]. In this architecture, each modality is first compressed by a two-layer MLP, producing modality-specific latent embeddings. These embeddings are then merged into a single shared representation via mean pooling. Integrating in latent space mitigates limitations of simple concatenation, including mismatched feature dimensionalities, varying feature scales, and the inability to capture higher-order cross-modal interactions. The multimodal encoder was implemented in Python with ReLU activations, dropout, the Adam optimiser, and a cross-entropy loss.

### Performance and Evaluation

F1 accuracy was the performance metric used for comparison to compensate for the three to one class imbalance in the AoU cohorts. We used nested cross-validation with three folds for performance estimation. We report the mean and standard deviation of accuracy across the three test folds. For each model, the splits were kept identical across all omics configurations and their combinations, ensuring that every data transformation was trained, validated, and tested on the same patients. For comparison, a stratified dummy predictor was trained as a baseline model.

For each multi-omics model, we performed paired significance tests comparing the performance of each transformed genetic feature set against the raw SNPs matrix. P-values were adjusted for multiple comparisons using the Benjamini-Hochberg correction across all transformations, models, cohorts, and diseases. Transformations that significantly outperformed the raw SNP matrix within the same model after correction are annotated with an asterisk (*).

## Results

### Individual Omic Predictions

As shown in Figure2, transcriptomics, proteomics, and metabolomics each substantially outperform genomics when considered in isolation. Proteomics alone, for example, achieves an F1 score of 0.75 *±*0.02 for CKD prediction, while no genomic transformation surpasses the dummy baseline in the AoU cohort. The PRS transformation is the only genomic modality to outperform any individual omic, achieving a modest F1 score of 0.58 *±* 0.02 for CKD prediction in the SHCS (Figure3 D). Taken together, these results highlight the limited standalone predictive power of genomics and motivate its integration with other omics for improved disease prediction.

**Figure 2.**
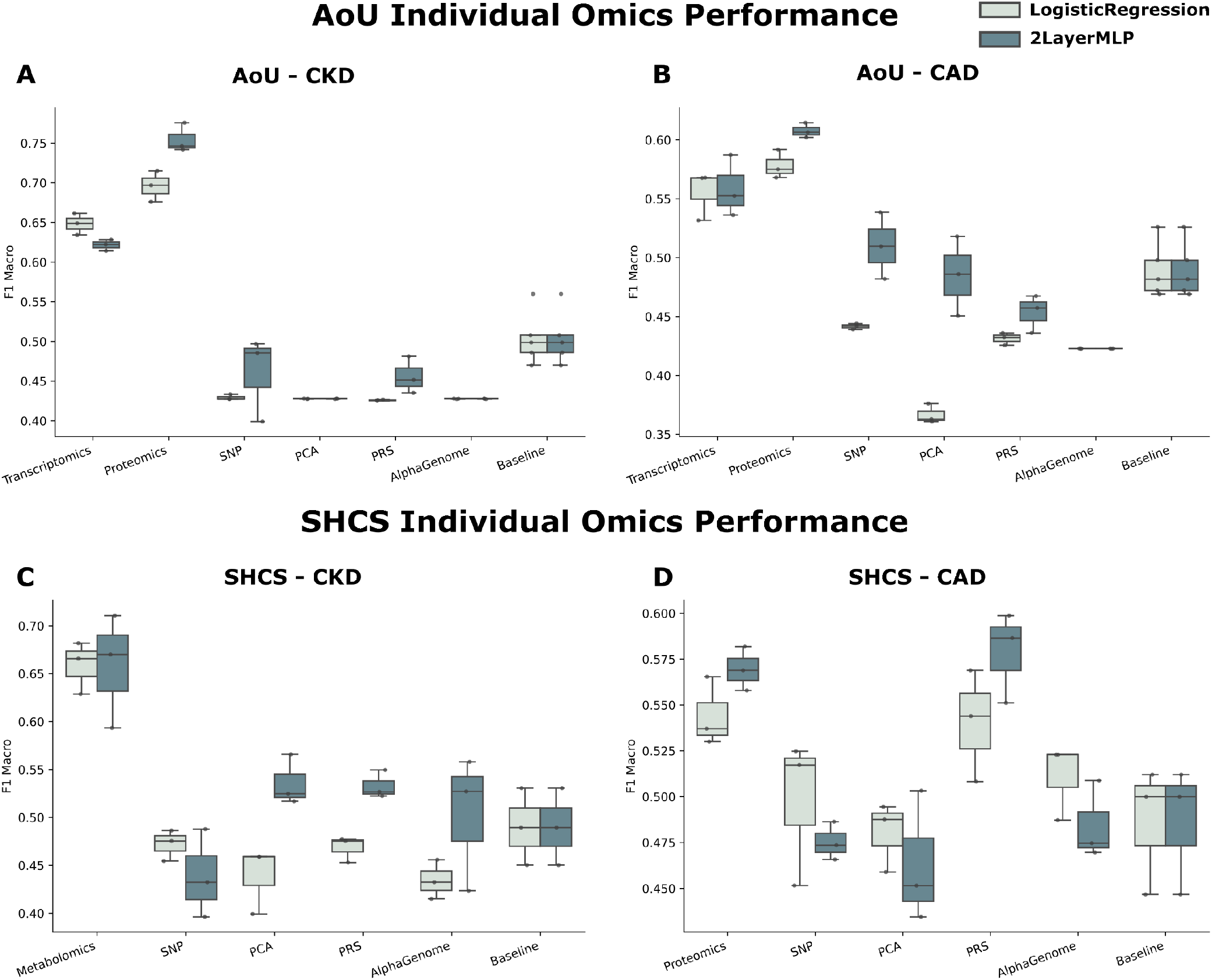
Performance of individual omics modalities for disease classification. Boxplots show macro-averaged F1 scores for logistic regression and a two-layer MLP using individual omics modalities, genetic feature representations and baseline predictors. Results are shown for CKD and CAD classification in the AoU (A, B) and the SHCS cohort (C, D), respectively. Boxes represent the interquartile range, centre lines indicate medians and whiskers show the observed variation across evaluation runs.

**Figure 3.**
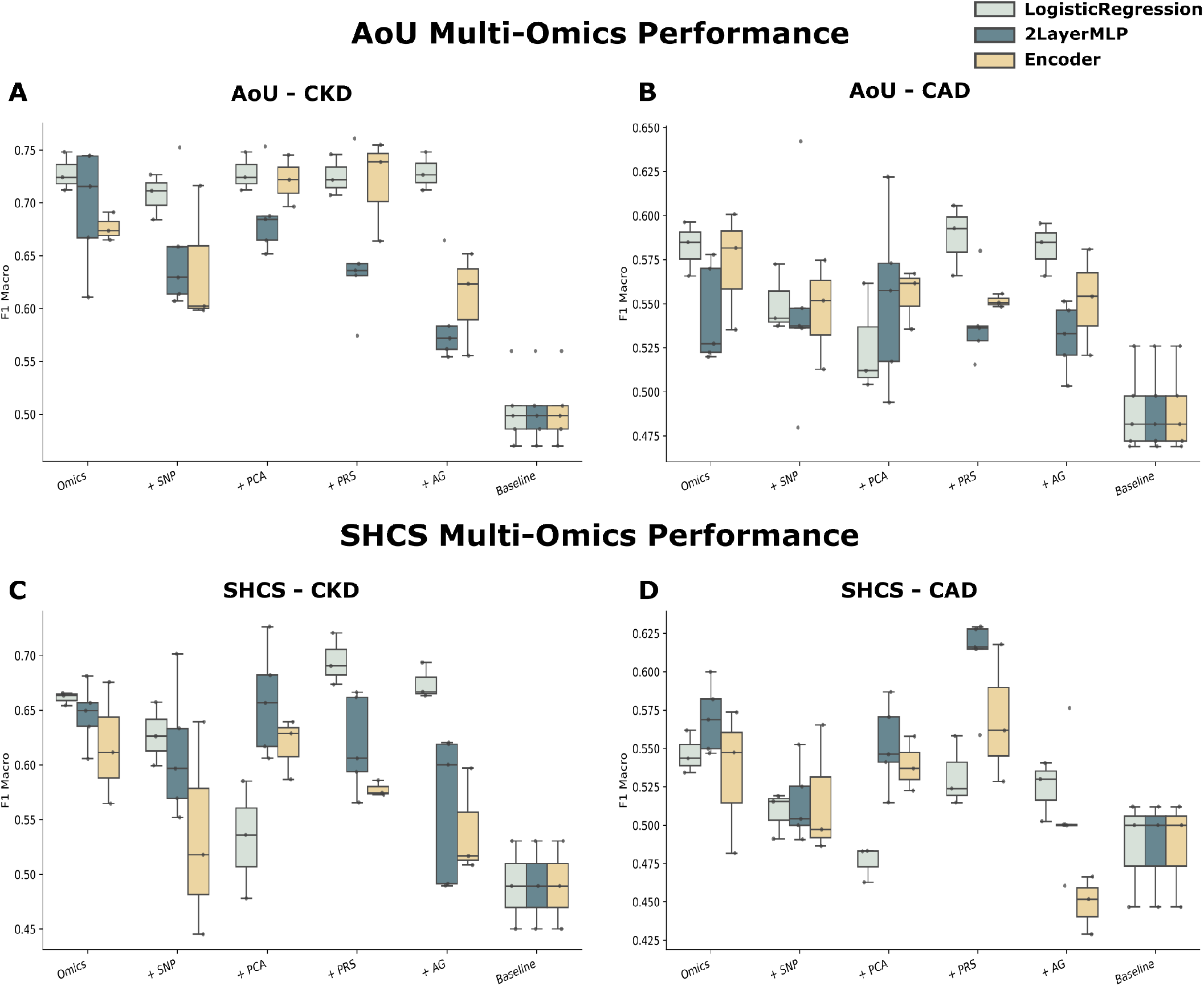
Performance of multi-omics for disease classification. Boxplots show macro-averaged F1 scores for logistic regression, a two-layer MLP, and a deep learning encoder using multi-omics modalities integrated with different genetic feature representations and a baseline predictor. Results are shown for CKD and CAD classification in the AoU (A, B) and the SHCS cohort (C, D), respectively. Boxes represent the interquartile range, centre lines indicate medians and whiskers show the observed variation across evaluation runs.

### Multi-Omics Predictions

The addition of genomics to other omics yields modest and variable gains, compared to individual omics predictions, depending on the choice of transformation. As shown in Figure 3, PRS was the most competitive transformation in the majority of cohort–disease combinations, matching or outperforming the multi-omics model alone and outperforming other genomic transformations in most cases. As per Table 2, it was statistically significantly higher at predicting CAD in the SHCS, even after adjusting for multiple corrections (FDR = 0.0053).

**Table 2.** MLP concatenation predictive performance.

| Transformation | AoU-CKD | AoU-CAD | SHCS-CKD | SHCS-CAD |
| --- | --- | --- | --- | --- |
| + SNP | $0.65 \pm 0.06$ | $0.55 \pm 0.06$ | $0.61 \pm 0.06$ | $0.51 \pm 0.02$ |
| + PCA | $0.69 \pm 0.04$ | $0.55 \pm 0.05$ | $0.66 \pm 0.05$ | $0.55 \pm 0.03$ |
| + PRS | $0.65 \pm 0.07$ | $0.54 \pm 0.02$ | $0.62 \pm 0.04$ | $0.61 \pm 0.03^*$ |
| + AlphaGenome | $0.59 \pm 0.04$ | $0.53 \pm 0.02$ | $0.56 \pm 0.07$ | $0.51 \pm 0.04$ |

Conversely, raw SNP matrices and PCA-based representations were consistently the weakest-performing transformations and, in several cases, degraded performance relative to the multi-omics model without genomics.

There was no clear optimal model architecture, however, in general the more complex encoder architectures did not improve upon simple feature concatenation. This could imply that the information contributed by genomic transformations is largely additive, and therefore better captured by simpler integration strategies.

## Discussion

Cross-modal interactions are crucial for capturing complementary information in multi-omics predictive modelling. However, genomic data remains largely underutilised or misrepresented in this context. This is partly due to its inherent complexity. Genomic data is non-continuous, sparse, and requires large cohorts to identify meaningful features. In this study, we evaluate a set of genomic data transformations that leverage pre-existing bioinformatic pipelines to determine whether alternative representations of genomic data can more effectively capture cross-modal interactions in multi-omics predictive modelling.

Across both the AoU and SHCS datasets, PRS emerged as the optimal transformation for genomic data integration in multi-omics predictive modelling. In the SHCS dataset, multi-omics models incorporating PRS achieved the highest F1 scores, outperforming all other genomic transformations, dummy baseline models, and unimodal omics models, highlighting the added value that PRS representations can bring to genomic integration. In the AoU dataset, no genomic representation improved performance beyond the joint proteomics and transcriptomics baseline; nevertheless, PRS did not degrade F1 performance. In contrast, both SNP matrix and PCA representations led to performance drops across CAD and CKD, highlighting the potential negative impact of naively incorporating these representations into a multi-omics predictive model.

No single integration method consistently outperformed the others. Notably, more complex encoder architectures failed to improve upon simple concatenation, suggesting that the genomic signals, as represented in this study, contribute in a linearly additive rather than a complementary manner. This contradicts what is known about genomic biology. Genomics forms intricate interactions with other omics layers. However, the complexity of these interactions likely far exceeds what can be captured given the sample sizes available in this study and the relatively compact deep learning models employed.

This also provides an intuitive explanation for why PRS is particularly well-suited to linear concatenation. Since PRS is itself a prediction, incorporating it via linear concatenation is functionally equivalent to ensemble prediction. This suggests that genomic features represented as PRS are best integrated through linear concatenation and, ideally, as a late integration strategy rather than the early integration approach employed here.

The complexity of genomics interactions further motivate adopting pipelines that integrate genomic data without requiring large sample sizes or high statistical power. In this study, we evaluated two such approaches, PRSs and AlphaGenome. By transforming unpruned SNPs into a lower-dimensional feature space, these methods preserve biological variability while incorporating known signal from GWAS summary statistics or from deep learning DNA foundation models. Such transformations will become increasingly important, as the cost of generating multiple omics remains high and multi-omics datasets rarely match genomics in scale. Therefore, identifying and benchmarking methods that integrate genomics without dampening predictive signal is of significant importance.

Future work should further explore AlphaGenome’s capabilities. A key advantage is its ability to capture regulatory and gene-expression signatures within ±1 Mb of a variant. In this study, we used the minimum window (16 kb) to focus on local effects on expression, however, future work could systematically vary the window size to assess trade-offs between locality and signal capture. AlphaGenome can also make predictions across tissues and molecular layers. Here, we scored variant effects on transcripts relevant to the comorbidities, but alternative tracks (e.g., DNase-seq, ATAC-seq) and multi-track, multi-tissue integration should be evaluated.

Overall, this study highlights the pitfalls of directly integrating genomics in multi-omics analyses and shows that applying bioinformatics transformations, such as PRS, improves performance. Future work should extend these findings in cohorts where both genomics and other omics are strongly informative, improve the AlphaGenome workflow, and benchmark more complex methods that optimally combine complementary signals. Overall, in this setting, models that combined other omics with PRS achieved the best accuracy for making clinical predictions in multi-omics integrative models.

## Data Availability

Deidentified individual participant data used in the study can be made available for investigators upon request to the corresponding author.

## Contributors

BR, CT, and JF contributed to study design. BR performed the predictive analyses. BR, MAO, and CT, performed data processing and transformations. BR, CT, and JF verified the underlying data and drafted the manuscript. All authors contributed to critical review and revision of the manuscript. CT and JF had final responsibility for the decision to submit for publication.

## Data sharing

Deidentified SHCS individual participant data used in the study can be made available for investigators upon request to the corresponding author.

All of Us data used in this study are available to approved researchers through the All of Us Research Program Researcher Workbench. Access can be requested at https://www.researchallofus.org.

## Code availability

All code is available at : https://github.com/Barry8197/GenDataTransformationsCode.

## Acknowledgements

We gratefully acknowledge All of Us participants for their contributions, without whom this research would not have been possible. We also thank the National Institutes of Health’s All of Us Research Program for making available the participant data and samples examined in this study.

This study was supported by the Swiss HIV Cohort Study (SHCS; project 960). SHCS data are gathered by the five Swiss university hospitals, two Cantonal hospitals, 15 affiliated hospitals and 36 private physicians (listed in http://www.shcs.ch/180-health-care-providers). The authors acknowledge the effort and commitment of SHCS participants, investigators, study nurses, laboratory personnel, and administrative assistance by the SHCS coordination and data centre.

## Notes

### Competing Interest Statement

The authors have declared no competing interest.

### Author Declarations

The SHCS was approved by the ethics committees of the participating institutions, BASEC-Nr. 2023-02080: Kantonale Ethikkommission Zurich; Ethikkommission Nordwest-und Zentralschweiz EKNZ; Kantonale Ethikkommission Bern; Commission Cantonale d'ethique de la recherche sur l'etre humain CCER-GE; Commission cantonale d'ethique de la recherche sur l'etre humain, CER-VD; Comitato etico cantonale, Ticino; Ethikkommission Ostschweiz EKOS, and written informed consent was obtained from all participants

### Summary of Updates

Included All of Us dataset as a validation dataset

## References

Z. Avsec, V. Agarwal, D. Visentin, J. R. Ledsam, A. Grabska-Barwinska, K. R. Taylor, Y. Assael, J. Jumper, P. Kohli, and D. R. Kelley. Effective gene expression prediction from sequence by integrating long-range interactions. Nature Methods, 18(10):1196–1203, Oct. 2021. ISSN 1548-7105. doi: 10.1038/s41592-021-01252-x. URL https://www.nature.com/articles/s41592-021-01252-x.

Z. Avsec, N. Latysheva, J. Cheng, G. Novati, K. R. Taylor, T. Ward, C. Bycroft, L. Nicolaisen, E. Arvaniti, J. Pan, R. Thomas, V. Dutordoir, M. Perino, S. De, A. Karollus, A. Gayoso, T. Sargeant, A. Mottram, L. H. Wong, P. Drotár, A. Kosiorek, A. Senior, R. Tanburn, T. Applebaum, S. Basu, D. Hassabis, and P. Kohli. Advancing regulatory variant effect prediction with AlphaGenome. Nature, 649(8099): 1206–1218, Jan. 2026. ISSN 1476-4687. doi: 10.1038/s41586-025-10014-0. URL https://www.nature.com/articles/s41586-025-10014-0.

A. Bonjoch, B. Bayés, J. Riba, J. Puig, C. Estany, N. Perez-Alvarez, B. Clotet, and E. Negredo. Validation of estimated renal function measurements compared with the isotopic glomerular filtration rate in an HIV-infected cohort. Antiviral Research, 88(3):347–354, Dec. 2010. ISSN 0166-3542. doi: 10.1016/j.antiviral.2010.09.015. URL https://www.sciencedirect.com/science/article/pii/S0166354210007370.

C. C. Chang, C. C. Chow, L. C. Tellier, S. Vattikuti, S. M. Purcell, and J. J. Lee. Second-generation PLINK: rising to the challenge of larger and richer datasets. GigaScience, 4 (1):s13742.#x2013;015–0047–8, Dec. 2015. ISSN 2047-217X. doi: 10.1186/s13742-015-0047-8. URL https://doi.org/10.1186/s13742-015-0047-8.

M. P. Cristelli, F. Cofán, N. Rico, J. C. Trullàs, C. Manzardo, F. Aguëro, J. L. Bedini, A. Moreno, F. Oppenheimer, J. M. Miro, F. Dieckman, A. Cases, E. Poch, E. Martinez, J. L. Blanco, F. García, J. Mallolas, J. M. Gatell, and the CKD-H. Clinic Investigators. Estimation of renal function by CKD-EPI versus MDRD in a cohort of HIV-infected patients: a cross-sectional analysis. BMC Nephrology, 18(1):58, Feb. 2017. ISSN 1471-2369. doi: 10.1186/s12882-017-0470-4. URL https://doi.org/10.1186/s12882-017-0470-4.

A. Gagneux-Brunon, P. Delanaye, N. Maillard, A. Fresard, T. Basset, E. Alamartine, F. Lucht, H. Pottel, and C. Mariat. Performance of creatinine and cystatin C-based glomerular filtration rate estimating equations in a European HIV-positive cohort. AIDS, 27(10):1573, June 2013. ISSN 0269-9370. doi: 10.1097/QAD.0b013e32835fac30. URL https://journals.lww.com/aidsonline/abstract/2013/06190/performance_of_creatinine_and_cystatin_c_based.6.aspx.

A. Khan, M. C. Turchin, A. Patki, V. Srinivasasainagendra, N. Shang, R. Nadukuru, A. C. Jones, E. Malolepsza, O. Dikilitas, I. J. Kullo, D. J. Schaid, E. Karlson, T. Ge, J. B. Meigs, J. W. Smoller, C. Lange, D. R. Crosslin, G. P. Jarvik, P. K. Bhatraju, J. N. Hellwege, P. Chandler, L. R. Torvik, A. Fedotov, C. Liu, C. Kachulis, N. Lennon, N. S. Abul-Husn, J. H. Cho, I. Ionita-Laza, A. G. Gharavi, W. K. Chung, G. Hripcsak, C. Weng, G. Nadkarni, M. R. Irvin, H. K. Tiwari, E. E. Kenny, N. A. Limdi, and K. Kiryluk. Genome-wide polygenic score to predict chronic kidney disease across ancestries. Nature Medicine, 28 (7):1412–1420, July 2022. 1038/s41591-022-01869-1. ISSN 1546-170X. doi: 10.1038/s41591-022-01869-1. URL https://www.nature.com/articles/s41591-022-01869-1.

S. A. Lambert, L. Gil, S. Jupp, S. C. Ritchie, Y. Xu, A. Buniello, A. McMahon, G. Abraham, M. Chapman, H. Parkinson, J. Danesh, J. A. L. MacArthur, and M. Inouye. The Polygenic Score Catalog as an open database for reproducibility and systematic evaluation. Nature Genetics, 53(4):420–425, Apr. 2021. ISSN 1546-1718. doi: 10.1038/s41588-021-00783-5. URL https://www.nature.com/articles/s41588-021-00783-5.

A. S. Levey, L. A. Stevens, C. H. Schmid, Y. L. Zhang, A. F. Castro, H. I. Feldman, J. W. Kusek, P. Eggers, F. Van Lente, T. Greene, and J. Coresh. A New Equation to Estimate Glomerular Filtration Rate. Annals of internal medicine, 150(9):604–612, May 2009. ISSN 0003-4819. doi: 10.7326/0003-4819-150-9-200905050-00006. URL https://pmc.ncbi.nlm.nih.gov/articles/PMC2763564/.

X. Li, J. Ma, L. Leng, M. Han, M. Li, F. He, and Y. Zhu. MoGCN: A Multi-Omics Integration Method Based on Graph Convolutional Network for Cancer Subtype Analysis. Frontiers in Genetics, 13, Feb. 2022. ISSN 1664-8021. doi: 10.3389/fgene.2022.806842. URL https://www.frontiersin.org/journals/genetics/articles/10.3389/fgene.2022.806842/full.

S. Ma, A. G. X. Zeng, B. Haibe-Kains, A. Goldenberg, J. E. Dick, and B. Wang. Moving towards genome-wide data integration for patient stratification with Integrate Any Omics. Nature Machine Intelligence, 7(1):29–42, Jan. 2025. s42256-024-00942-3. ISSN 2522-5839. doi: 10.1038/ URL https://www.nature.com/articles/s42256-024-00942-3.

A. Mocroft, J. D. Lundgren, M. Ross, C. A. Fux, P. Reiss, O. Moranne, P. Morlat, A. d. Monforte, O. Kirk, and L. Ryom. Cumulative and current exposure to potentially nephrotoxic antiretrovirals and development of chronic kidney disease in HIV-positive individuals with a normal baseline estimated glomerular filtration rate: a prospective international cohort study. The Lancet HIV, 3(1):e23–e32, Jan. 2016. ISSN 2352-3018. doi: 10.1016/S2352-3018(15)00211-8. URL https://www.sciencedirect.com/science/article/pii/S2352301815002118.

P. Patel, M. Wang, Y. Ruan, S. Koyama, S. L. Clarke, X. Yang, C. Tcheandjieu, S. Agrawal, A. C. Fahed, P. T. Ellinor, P. S. Tsao, Y. V. Sun, K. Cho, P. W. F. Wilson, T. L. Assimes, D. A. van Heel, A. S. Butterworth, K. G. Aragam, P. Natarajan, and A. V. Khera. A multi-ancestry polygenic risk score improves risk prediction for coronary artery disease. Nature Medicine, 29(7):1793–1803, July 2023. ISSN 1546-170X. doi: 10.1038/s41591-023-02429-x. URL https://www.nature.com/articles/s41591-023-02429-x.

Ryan, R. E. Marioni, and T. I. Simpson. MultiOmic Graph Diagnosis (MOGDx): a data integration tool to perform classification tasks for heterogeneous diseases. Bioinformatics, 40(9):btae523, Sept. 2024. ISSN 1367-4811. doi: 10.1093/bioinformatics/btae523. URL https://doi.org/10.1093/bioinformatics/btae523.

I. C. Schoepf, C. W. Thorball, B. Ledergerber, T. Engel, M. Raffenberg, N. A. Kootstra, P. Reiss, B. Hasse, C. Marzolini, C. Thurnheer, M. Seneghini, E. Bernasconi, M. Cavassini, H. Buvelot, R. Kouyos, H. F. Günthard, J. Fellay, P. E. Tarr, and Swiss HIV Cohort Study. Coronary Artery Disease–Associated and Longevity-Associated Polygenic Risk Scores for Prediction of Coronary Artery Disease Events in Persons Living With Human Immunodeficiency Virus: The Swiss HIV Cohort Study. Clinical Infectious Diseases, 73(9):1597–1604, Nov. 2021. ISSN 1058-4838. doi: https://doi.org/10.1093/cid/ciab521. URL 10.1093/cid/ciab521.

T. Wang, W. Shao, Z. Huang, H. Tang, J. Zhang, Z. Ding, and K. Huang. MOGONET integrates multi-omics data using graph convolutional networks allowing patient classification and biomarker identification. Nature Communications, 12 (1):3445, June 2021. ISSN 2041-1723. doi: 10.1038/s41467-021-23774-w. URL https://www.nature.com/articles/s41467-021-23774-w.

